# Knowledge, Attitudes, and Practices Regarding Antimicrobial Resistance Among Users of an Indian Digital Healthcare Platform: A Cross-sectional Online Survey

**DOI:** 10.64898/2026.10.14.26365518

**Authors:** Zoya Agarwal, Swati Mishra, Jasmine Kaur, Rajeev Sharma, Tavpritesh Sethi

## Abstract

**Background:** Antimicrobial resistance (AMR) is a growing global health threat, and India’s high antibiotic consumption makes public knowledge, attitudes, and practices (KAP) a critical determinant of stewardship outcomes. Existing Indian KAP evidence is drawn from small, localised cohorts, leaving a gap in nationwide, digitally-sourced data. This study aimed to describe antibiotic-related knowledge, attitudes and self-reported practices among users of a digital healthcare platform in India, examine demographic and regional variation, and identify potential areas for future stewardship interventions.

**Methods:** A cross-sectional survey was conducted among adult users of the Tata 1mg platform (December 2025–February 2026), with participants invited via an in-app banner. Knowledge and attitude were each scored on a 0-4 scale (4 items); practice was scored as a percentage of applicable items answered favourably, with scoring adjusted for branching logic based on antibiotic-access route. Of 10,726 respondents who started the survey, 4,883 completed it (45.5%). City of residence was mapped to one of six regions (India’s five statutory Zonal Council zones plus a North-Eastern region) for 4,187 respondents. Associations with demographic characteristics were assessed using multivariable linear regression, and regional differences were assessed using one-way ANOVA. A parallel survey of 256 physicians was conducted to enable patient–provider comparison.

**Results:** Mean scores were: knowledge 2.28 (SD 1.19), attitude 0.91 (SD 1.14), practice 44.9% (SD 33.4); attitude, not knowledge, was the weakest domain. Older age was associated with higher scores across all domains (practice: B=12.52, p<0.001 for age ≥65 vs. 18-34); city tier was independently associated with lower knowledge and attitude (Tier 3 vs. Tier 1: p=0.010 and p<0.001, respectively). Regional differences were non-significant for knowledge and practice but significant for attitude (F=2.56, p=0.026, η^2^=0.003), driven by lower scores in the North-Eastern region. In the parallel physician survey, 55.5% of respondents estimated that more than half of their patients self-medicated before consultation.

**Conclusion:** In this platform-based sample, attitudes were the weakest domain, not knowledge, and 42.0% said their last antibiotic did not come on a doctor’s recommendation. The findings may inform digital stewardship touchpoints, such as point-of-purchase guidance and prescription verification, in line with India’s National Action Plan on Antimicrobial Resistance 2.0. As a cross-sectional survey of platform users, it cannot establish the relative contributions of knowledge, beliefs and access pathways to antibiotic use.

## Introduction

Antimicrobial resistance (AMR) has emerged as a critical public health challenge. Globally, one in six laboratory-confirmed bacterial infections is now resistant to standard antibiotics, a figure that rises to approximately one in three in the WHO South-East Asia region [1]. According to recent Global Research on Antimicrobial Resistance (GRAM) Project estimates, bacterial AMR was linked to nearly 5 million deaths, with 1.27 million directly caused by it [2]. Without major shifts in how we use antibiotics, this burden will continue to grow [3]. In fact, resistance rose across more than 40% of monitored pathogen-antibiotic combinations between 2018 and 2023 [1]. Alarmingly, AMR continues to outpace medical innovation, with roughly 40 years having passed since the discovery of the last genuinely new antibiotic class [4]. India, historically one of the world’s largest consumers of antibiotics[5], faces a substantial and evolving burden of AMR driven by a complex interplay of high-risk behaviours and structural access pathways, such as easy over-the-counter availability and self-medication[6, 7].

Existing literature on AMR in India has predominantly documented a persistent awareness-practice paradox where high levels of self-reported awareness among patients fail to mitigate high-risk behaviors such as self-medication. A large multicentric community KAP survey conducted across India reported that while participants demonstrated appropriate knowledge and attitudes toward antibiotic use to some degree, important knowledge gaps and weak self-medication practices persisted across the sampled populations[8]. Similarly, a community-based cross-sectional study among low-income households in urban Delhi found that although a majority of residents had heard of antibiotics, specific knowledge of appropriate use and of the mechanisms underlying resistance remained poor, and self-medication with leftover or shared antibiotics was common[9]. Another cross-sectional survey of the general public in coastal south Karnataka reported comparable gaps in the recognition of antibiotic misuse and continued reliance on non-prescription sources[10], while a study among the population attending a large mass-gathering event in central India similarly found that awareness of AMR terminology did not protect against self-treatment of upper respiratory tract infections with antibiotics[11]. However, this evidence base has historically relied mostly on localised, small-scale clinical, community, or event-based cohorts, typically ranging from a few hundred to around two thousand participants drawn from a single city or region.

Current literature presents two crucial limitations. Primarily, it lacks a nationwide baseline regarding how these knowledge-practice disconnects manifest at a geographically diverse, pan-India scale. Secondly, there is a distinct shortage of data within the new, evolving digital ecosystem. Digital healthcare platforms are becoming an increasingly important point of contact between the Indian public and antibiotics, through ePharmacy, teleconsultation, and eDiagnostics. Yet, to our knowledge, no extensive KAP survey has been conducted within such an ecosystem among the growing population of users who access healthcare through digital platforms rather than through traditional in-person channels. Establishing a comprehensive baseline through a contemporary channel is essential for evolving national policy, specifically the National Action Plan on Antimicrobial Resistance 2.0 (NAP-AMR 2.0 2025-2029). This approach involving digital healthcare platforms, with their expanding reach and frequent interaction with patients, offer a valuable opportunity to deliver customized stewardship interventions at scale, tailored to both patients and physicians.

To address the limited availability of large-scale evidence from digitally engaged populations, we conducted a large-scale cross-sectional survey of 4,883 adult users of a national digital health platform in India. To our knowledge, this is among the largest platform-based assessments of antibiotic-related knowledge, beliefs, and self-reported practices conducted in the country. We examined variation across demographic characteristics, city tiers, and geographic zones and, as an exploratory secondary analysis, compared selected public responses with conceptually related findings from a parallel survey of practising physicians. The study was additionally intended to identify actionable targets for future digital antimicrobial-stewardship interventions.

## Materials and Methods

### Study design and setting

A cross-sectional, online Knowledge, Attitudes and Practices (KAP) survey was conducted among adult users of the Tata 1mg platform over a 12-week period from December 2025 to February 2026. Ethics approval and consent are described under Declarations.

### Study population

The study population comprised registered Tata 1mg users, aged 18 years or older residing in India. Individuals below 18 years are unable to register on the platform because of application registration controls and app-store eligibility restrictions. Tata 1mg is one of India’s largest digital healthcare platforms, serving over 260 million registered users across more than 1,800 cities through services including online pharmacy, diagnostics, teleconsultation, and chronic care management. Its nationwide reach provided an opportunity to capture responses from users across diverse geographic regions and urban settings.

Exclusion criteria were duplicate submissions, incomplete questionnaires (defined as responses that did not reach the final item), any malformed responses (for example, only a single logged answer with no other data, consistent with a data-logging artefact rather than genuine survey engagement).

Participants were selected using a non-probability convenience sampling approach enabling participation from users across different regions of India and a range of urban settings.

### Sample size estimation

The sample size was estimated using Cochran’s formula:

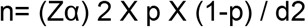

where n is the required sample size, Zα is the standard normal deviate corresponding to the desired confidence level (1.96 for 95% confidence), p is the expected proportion (based on previous studies or pilot data), and d is the precision (allowable margin of error).

The sample size was calculated based on the primary objective of estimating the prevalence of self-medication with antibiotics without consulting a doctor.

Assuming a prevalence of 24.6% from prior studies[9], with a 95% confidence level (Z = 1.96) and 5% absolute precision, the initial sample size calculated using the Cochran formula was 285. This was further adjusted for a design effect of 2 to account for clustering, resulting in 570 participants. Considering an anticipated response rate of 20%, the final sample size required to be approached was 2850. To ensure adequate statistical power and account for potential exclusions, the target sample size was rounded to 3,000 participants[12].

### Questionnaire development

The survey questionnaire was developed by adapting selected items from the World Health Organization (WHO) multi-country public awareness survey on antibiotic use and antimicrobial resistance (2015)[13], supplemented with items newly developed for the Indian digital healthcare setting, within the conceptual domains of knowledge, attitudes, and practices regarding antibiotic use and AMR (Supplementary Table 1). It was administered in English, the primary language of the platform.

A modified and shortened instrument was developed to improve usability within an Indian mobile application environment. Lengthy online questionnaires are usually associated with reduced completion rates and increasing respondent fatigue. The final questionnaire comprised 16 items: 13 closed-ended items distributed across three KAP domains (practice, attitude, and knowledge), and 3 demographic variables, of which age and gender were closed-ended and city of residence was collected as free text. Questions were organised in the sequence of practice, attitude, and knowledge to minimise response priming and social desirability bias (Supplementary Table 2).

The practice section incorporated conditional branching to ensure that respondents received questions relevant to their most recent antibiotic use. Participants reporting that their last antibiotic had been recommended by a physician followed a different pathway from those who obtained antibiotics through non-physician sources. To support recall accuracy for practice-domain items and capture respondents’ most recent experiences, antibiotic use was evaluated within the preceding 12 months.

### Questionnaire validation

Prior to implementation, the questionnaire underwent face validation to assess its clarity, readability and overall comprehensibility among the intended target population. Feedback from this process informed certain revisions to question wording and response options to improve clarity without changing the underlying constructs. Further suggestions, namely a broader knowledge domain, an item on where respondents get information about antibiotics, and a version in languages other than English, would have required expanding the item pool, translation and re-piloting of the instrument, so they were held for a future version. The revised questionnaire was then finalised for use in the main study.

### Data collection

Data were collected digitally using **Targeted User Automated Surveys (TAUS)**, an in-house developed survey platform integrated within the Tata 1mg application. Participants were invited through the digital health platform’s customer relationship management (CRM) channels, including email and in-app notifications.

Survey responses were captured electronically and stored in anonymised form. Platform-based user identifiers restricted each participant to a single submission, thereby reducing the likelihood of duplicate responses. Response options were randomised for participants wherever appropriate to mitigate order bias.

### Variable definitions

The primary outcome measures were the domain-specific knowledge, attitude and practice scores derived from the questionnaire.

Age, gender and city tier were included as explanatory variables. Age was categorised into 18-34 years, 35-64 years and ≥65 years to reflect differences in healthcare utilisation patterns across the adult life course. The 18–34-year group served as the reference category for regression analyses. Gender was recorded as male, female or prefer not to mention. For the geographic mapping analysis described in Results, city or town of residence is based primarily on the respondent’s own free-text entry, supplemented by a platform-derived location field where that entry was missing or incomplete. These were additionally mapped to Indian states, followed by categorization under one of six regions: the five statutory zones established under the Zonal Councils framework of the States Reorganisation Act, 1956, and maintained by the Ministry of Home Affairs[14], plus a sixth North-Eastern region defined under the North Eastern Council Act, 1971 (including Sikkim)[15]. Additionally, the city list was mapped to a three-tier hierarchy using the Government of India, Department of Expenditure Office Memorandum on classification of cities (7th Central Pay Commission, 2015): Tier 1 corresponds to the eight Government-designated ‘X’ cities, Tier 2 to the Government-designated ‘Y’ cities, and Tier 3 to all remaining (‘Z’/unclassified) cities and towns[16].

Information on education and household income was intentionally not collected to minimise respondent burden and reduce the potential for self-reporting and social desirability bias in the digital survey environment.

### Questionnaire scoring

Questions relating to age, gender and city of residence were collected for descriptive purposes and were not included in the scoring. Similarly, the question assessing antibiotic use during the previous 12 months functioned as a self-reported exposure and stratification variable.

The questionnaire generated three domain-specific scores comprising knowledge (0-4) and attitude (0-4). The practice domain used branching logic: respondents who selected “Doctor” were routed directly to Questions 7 and 8, while all other responses were routed to Questions 6 and 7. As a result, the number of applicable practice items varied between 2 and 3 depending on the respondent’s care pathway. For all scored items, correct responses received one point, whereas incorrect, “Not sure” and “Don’t know/Don’t remember” responses received zero points (Supplementary Tables 2 & 3). For the multiple-response question assessing misuse of antibiotics, a score of one was awarded if respondents identified at least one correct misuse practice, including instances where all correct options were selected. Higher scores indicated better knowledge, more appropriate attitudes and more appropriate antibiotic-use practices.

#### Parallel physician survey

A shorter KAP questionnaire was concurrently circulated to physicians across India through professional networks and messaging groups to contextualise the general public-reported findings against a physician-side perspective. Of 779 responses received, 256 were fully completed and form the analytic sample reported under Results (*Comparative Highlights: Patients versus Physicians*).

The physician survey instrument consisted of 20 items, including 4 demographic and professional background variables, 7 knowledge items, 4 attitude items, 4 practice items, and 1 open-ended question on challenges in antibiotic use asking their opinions, to capture contextual insights. The questions and scoring rationale for each item is presented alongside the scores in Supplementary Table 4.

> ***Given the scope of the present study, physicians’ knowledge, attitude, and practice findings were not evaluated as a standalone KAP assessment in their own right. The physician data were instead restricted to a set of items comparable across both surveys, used solely to triangulate against the corresponding patient-reported findings***.

### Statistical analysis

Only completed questionnaires were included in the final analytical dataset for both the general-public and the physician surveys, no imputation for missing data was required. A response was classified as complete if all knowledge, attitude, and practice items were answered; demographic items (age, gender, and city of residence) were not required for a response to be classified as complete, and their completion is reported separately in Table 1.

**Table 1:** Socio-demographic characteristics of respondents (N = 4,883)

| Characteristic | Category | n | % |
| --- | --- | --- | --- |
| **Age group** | 18–34 years | 2,915 | 59.7 |
|  | 35–64 years | 1,560 | 31.9 |
|  | ≥65 years | 408 | 8.4 |
| **Gender** | Male | 3,202 | 65.6 |
|  | Female | 1,621 | 33.2 |
|  | Prefer not to mention | 60 | 1.2 |
| **City tier (DoE, 2015 X/Y classification)\*** | Tier 1 (X cities) | 1,289 | 30.7 |
|  | Tier 2 (Y cities) | 1,028 | 24.5 |
|  | Tier 3 (all other cities/towns) | 1,878 | 44.8 |
|  | Not applicable (location unverified) | 688 | - |
| **Antibiotic use in past 12 months (exposure variable)** | Yes | 2,190 | 44.8 |
|  | No | 1,883 | 38.6 |
|  | Don't remember | 810 | 16.6 |

*Note: City tier is reported for the 4,195 respondents whose reported location could be verified against a known Indian city, town, or district; a reliable tier could not be assigned for the remaining 688 respondents (14.1%). Age, gender, and antibiotic-use exposure are reported for the full analytic sample (N = 4,883), as these do not depend on location verification.*

All analyses were performed using anonymised datasets, and results are presented only in aggregate form. Knowledge and attitude are reported on their native 0-4 scale (equivalently, 57.0% and 22.7% of the maximum); practice domain is reported as a percentage because its denominator varies by branch.

Participant characteristics and questionnaire responses were summarised using descriptive statistics. Categorical variables are presented as frequencies and percentages, while continuous variables are summarised using means and standard deviations. Associations between participant characteristics and domain-specific knowledge, attitude and practice scores were examined using separate linear regression models. A *p*-value of <0.05 was considered statistically significant.

## Results

A total of 3,181,073 platform users were exposed to the study’s recruitment banner during the survey collection period. Of these, 10,782 opened the survey, and 10,726 went on to respond to at least the first item. Of those respondents, 4,883 completed the survey in full, a completion rate of 45.5% relative to those who started (not those who merely opened the survey or saw the banner). More than half of respondents (52.8%) progressed beyond the 10th of 16 items (Supplementary Feigure 1).

### Participant characteristics

Of the 4,883 completed respondents, the largest age group was 18-34 years (n = 2,915; 59.7%), followed by 35-64 years (n = 1,560; 31.9%) and 65 years and above (n = 408; 8.4%). The majority identified as male (n = 3,202; 65.6%), followed by female respondents (n = 1,621; 33.2%) and 1.2% preferring not to disclose gender (n = 60). Among the 4,195 respondents with a verified location, nearly half were from Tier 3 (n = 1,878; 44.8%), followed by Tier 1 (n = 1,289; 30.7%) and Tier 2 (n = 1,028; 24.5%). Table 1 summarises the socio-demographic profile of the analytic sample.

#### Findings from the Knowledge section

The mean knowledge score was 2.28 (SD 1.19) out of a maximum of 4, indicating that, on average, respondents answered just over half of the knowledge items correctly. Recognition of behaviours constituting antibiotic misuse was the best-performing item (78.1% correct), whereas fewer than half of respondents (42.3%) reported that a doctor had ever explained antibiotic resistance to them, and only about half correctly identified the core definition of AMR (51.3%) or affirmed it as a serious threat (56.9%). Table 2 presents the proportion of respondents giving the correct/knowledgeable response to each item.

**Table 2:** Pooled analysis of knowledge-domain items (N = 4,883)

| Knowledge item | Correct response | N answered | n correct | % correct |
| --- | --- | --- | --- | --- |
| Doctor has explained antibiotic resistance (AMR) | Yes | 4,883 | 2,064 | 42.3 |
| AMR is a growing, serious threat for all of us | True | 4,883 | 2,776 | 56.9 |
| AMR occurs when antibiotics fail to treat infections | True | 4,883 | 2,505 | 51.3 |
| Correctly identifies at least one true form of antibiotic misuse (skipping doses / stopping course early / taking without prescription / using leftovers) | ≥ 1 correct option selected | 4,883 | 3,815 | 78.1 |

#### Findings from the Attitudes section

The mean attitude score was 0.91 (SD 1.14) out of a maximum possible score of 4. Across the four attitude items, favourable responses ranged from 15.2% (disagreeing that antibiotics help resolve a cough or cold) to 31.7% (disagreeing that leftover antibiotics can be reused). Most respondents either agreed with or were unsure about items assessing potentially inappropriate beliefs about antibiotic use. In particular, 77.4% agreed with or were unsure that mild antibiotics can be used if recommended by a pharmacist, while 84.8% agreed with or were unsure that antibiotics help shorten the course of a cough or cold. Response distributions are presented in Table 3.

**Table 3:** Pooled analysis of attitude-domain items (N = 4,883)

| Attitude item | Favourable response | N answered | n favourable | % favourable |
| --- | --- | --- | --- | --- |
| Antibiotics are safe and effective, so they can be used commonly | Disagree | 4,883 | 1,045 | 21.4 |
| Taking antibiotics can stop a cough or cold from getting worse | Disagree | 4,883 | 740 | 15.2 |
| Some mild antibiotics are okay to be taken if a pharmacist recommends them | Disagree | 4,883 | 1,104 | 22.6 |
| Leftover or unused antibiotics can be taken for a similar illness in future | Disagree | 4,883 | 1,548 | 31.7 |

#### Findings from the Antibiotic-use practices section

By design, the diagnostic-test question was only administered to respondents who reported that their last antibiotic use had been recommended by a doctor. Since the number of eligible practice items depended on which branch a respondent fell into (2 or 3, depending on the “Doctor” versus non-”Doctor” path), Practice is shown as a percentage, rather than a raw score out of 4 like Knowledge and Attitude. The composite practice score was computed as the percentage of applicable items answered favourably-out of 3 items (healthcare-seeking behaviour, course completion, and diagnostic testing) for the doctor-recommended subgroup, and out of 2 items (healthcare-seeking behaviour and course completion) for the remaining subgroup. This approach ensured that each respondent’s score reflects performance only on the items relevant to their own care pathway, and is comparable in scale across both subgroups despite the difference in item count. On this metric, the mean practice score was 44.9% (SD 33.4). Just over half of respondents (58.0%) reported that their last antibiotic use followed a doctor’s recommendation, a similar proportion (46.7%) of those who saw a doctor reported that a diagnostic test had been performed before prescribing, and fewer than half of all respondents (42.1%) reported always completing the full antibiotic course (Table 4).

**Table 4:** Pooled analysis of practice-domain items (N = 2,833-4,883)

| Practice item | Favourable/correct response | N answered | n favourable | % favourable |
| --- | --- | --- | --- | --- |
| Healthcare-seeking behaviour for last antibiotic use (derived from Q5 branch) | Doctor recommended (Q5 = a) | 4,883 | 2,834 | 58.0 |
| Completed the full antibiotic course | Always | 4,883 | 2,058 | 42.1 |
| Doctor recommended a diagnostic test before prescribing (doctor-recommended subgroup only) | Yes | 2,833 | 1,324 | 46.7 |

**Note:**
Three practice items were assessed. Because Question 5 (source of the last antibiotic recommendation) branched respondents into two pathways — those reporting a doctor's recommendation (5a) proceeded to questions on course completion and diagnostic testing, while those reporting a pharmacist, self-medication, family/friend recommendation, or a repeated previous prescription (5b–5e) proceeded to a question on their reason for not consulting a doctor and a parallel course-completion item - the diagnostic-testing item was, by design, administered only to the doctor-recommended subgroup. Denominators therefore differ across items, as shown in Table 4.

This is the same approach we have followed for Practice throughout the other analyses in this paper.

#### Knowledge, attitude and practice by recent antibiotic-use status

Table 5 presents knowledge, attitude and practice scores by antibiotic use in the past 12 months (Q4 response). Mean scores were as follows: Knowledge 2.54 (Yes), 2.15 (No), 1.90 (Don’t remember); Attitude 1.06 (Yes), 0.82 (No), 0.71 (Don’t remember); Practice 50.7% (Yes), 41.0% (No), 38.3% (Don’t remember). One-way ANOVA showed statistically significant differences across the three groups for all three domains: Knowledge F = 110.10, p < 0.001; Attitude F = 39.92, p < 0.001; Practice F = 63.02, p < 0.001.

**Table 5:** Knowledge, attitude and practice scores by recent antibiotic-use status (N = 4,883)

| Antibiotic use in past 12 months | n | Knowledge, mean (SD) | Attitude, mean (SD) | Practice %, mean (SD) |
| --- | --- | --- | --- | --- |
| Yes | 2,190 | 2.54 (1.07) | 1.06 (1.19) | 50.7 (32.9) |
| No | 1,883 | 2.15 (1.24) | 0.82 (1.09) | 41.0 (33.1) |
| Don't remember | 810 | 1.90 (1.22) | 0.71 (1.02) | 38.3 (32.7) |

#### Domain scores by gender, age group, and city tier

Descriptively, knowledge, attitude, and practice scores were all lowest among the youngest age group (18-34 years) and highest among respondents aged 65 years and above. Scores were marginally higher among men than women, and were highest in Tier 1 cities, and generally lower in Tier 2 and Tier 3 (with knowledge scores nearly identical between the two). Table 6 presents mean domain scores stratified by gender, age group, and city tier. Knowledge and attitude scores are expressed on a 0-4 scale (higher = more knowledgeable / more favourable attitude); the practice score is expressed as a percentage (0-100%) of applicable items answered favourably. Summary of geographical mapping of cities is provided in Supplementary Table 5.

**Table 6:** Mean Domain scores stratified by gender, age group, and city tier (N = 4,883)

| Stratum | n | Knowledge, mean (SD) | Attitude, mean (SD) | Practice %, mean (SD) |
| --- | --- | --- | --- | --- |
| **Gender** |  |  |  |  |
| Male | 3,202 | 2.32 (1.19) | 0.95 (1.18) | 45.1 (33.5) |
| Female | 1,621 | 2.22 (1.18) | 0.83 (1.05) | 44.7 (33.2) |
| Prefer not to mention | 60 | 2.20 (1.16) | 1.05 (1.11) | 40.8 (34.8) |
| **Age group** |  |  |  |  |
| 18-34 years | 2,915 | 2.23 (1.18) | 0.80 (1.07) | 41.7 (33.4) |
| 35-64 years | 1,560 | 2.35 (1.20) | 1.02 (1.21) | 48.2 (32.8) |
| ≥65 years | 408 | 2.46 (1.14) | 1.26 (1.27) | 54.7 (32.5) |

| City tier* |  |  |  |  |
| --- | --- | --- | --- | --- |
| **Tier 1** | 1,289 | 2.37 (1.19) | 1.05 (1.21) | 47.0 (33.2) |
| **Tier 2** | 1,028 | 2.26 (1.20) | 0.97 (1.18) | 44.9 (33.4) |
| **Tier 3** | 1,878 | 2.26 (1.20) | 0.84 (1.12) | 44.7 (33.4) |

Note: \*City tier is reported for the 4,195 respondents with a verified location (see Table 1 note); gender and age group are reported for the full N = 4,883.

#### Associations between participant characteristics and domain scores

Older age was independently and consistently associated with higher scores across all three domains (all p<0.01 for the 35-64 and ≥65 categories relative to 18-34 years), with the largest effect seen for the practice score (12.5 percentage-point higher mean score in the ≥65 group). Females scored lower than males on attitude (B = −0.11, p = 0.005). The knowledge difference was smaller and not statistically significant (B = −0.075, p = 0.059), and practice was similar in the two groups (p = 0.280). Residing in a Tier 3 city was independently associated with lower scores on knowledge (p = 0.010) and attitude (p<0.001) relative to Tier 1, though not significantly for practice (p = 0.097); the Tier 2 vs. Tier 1 difference reached significance for knowledge only (p = 0.023). The proportion of variance explained by these demographic variables was modest for all three models (R^2^ = 0.007-0.026), indicating that gender, age, and city tier account for only a small fraction of the variability in AMR-related knowledge, attitudes, and practices (Table 7).

**Table 7:**
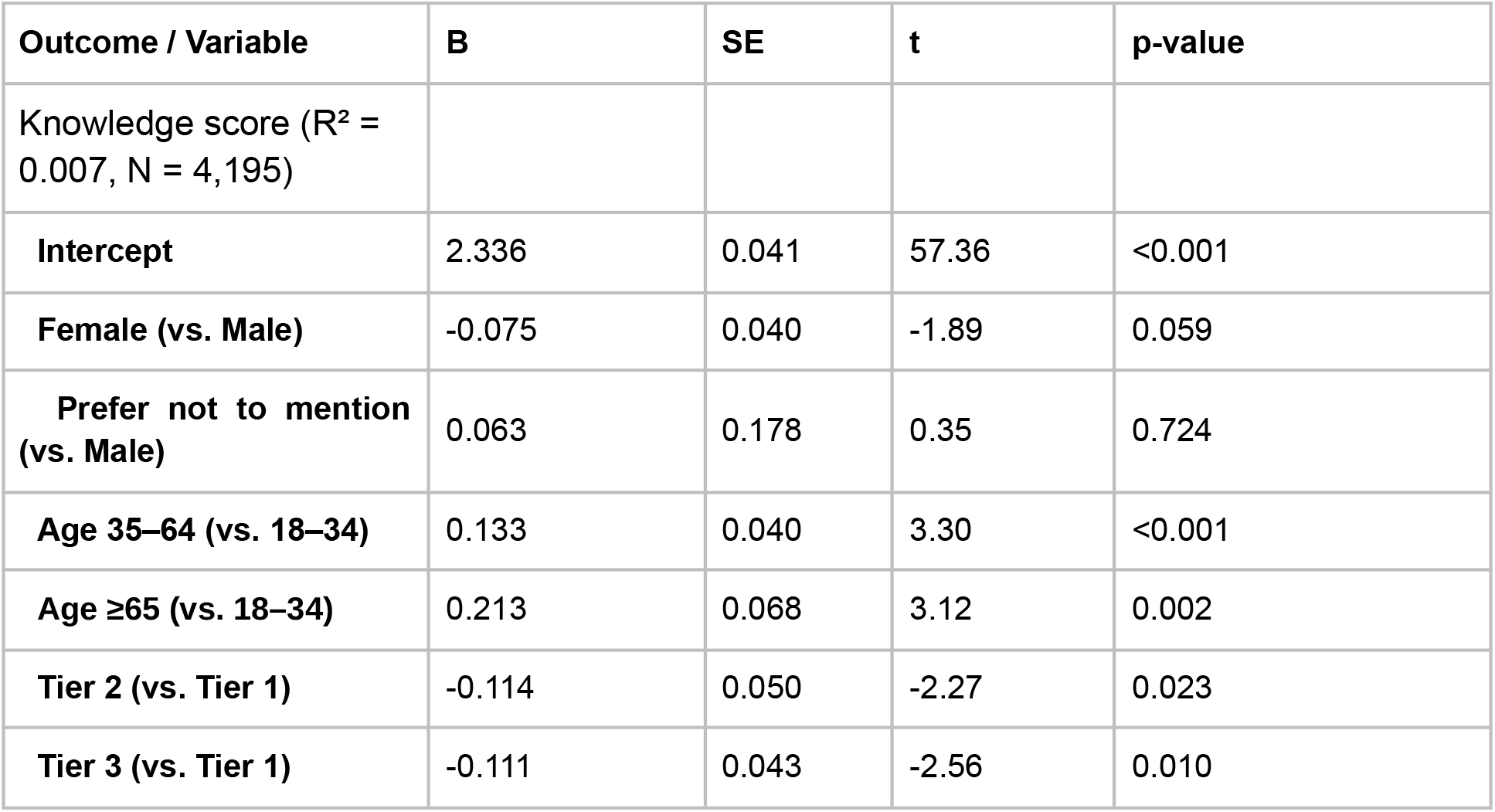
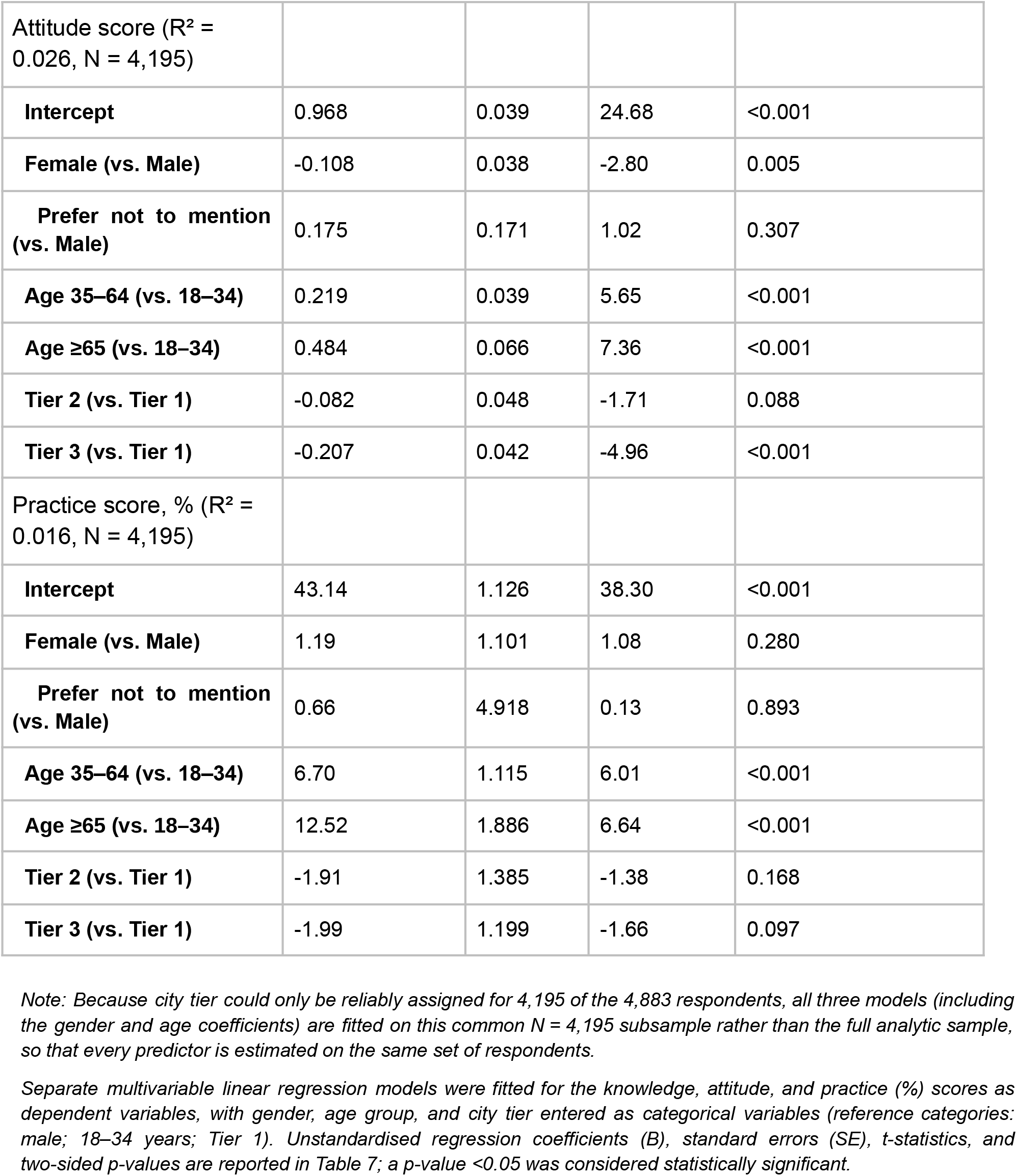
Linear regression of knowledge, attitude, and practice scores on gender, age group, and city tier (N = 4,195)

### Regional mapping and Geographic distribution

Of the 4,883 completed respondents, 4,195 (85.9%) had a city that could be reliably mapped to an Indian state. Of these, 4,187 fell within one of six regions: Northern, Central, Eastern, Western and Southern under the Zonal Councils framework[14], and North-Eastern under the North Eastern Council Act, 1971 (including Sikkim)[15]. The Union Territories of Delhi, Chandigarh, Puducherry, and Dadra and Nagar Haveli and Daman and Diu were each grouped with their respective zones under the Zonal Councils framework and, on this basis, placed within their corresponding regions. Only the Andaman and Nicobar Islands, which falls outside both the Zonal Councils framework and the North Eastern Council, was excluded from the six-region classification; respondents from this Union Territory (n = 8) are reported separately as unclassified. A state-wise breakdown of the number of respondents by region is provided in Supplementary Table 6. The mapped sample across the six regions comprised Northern (n = 903), Central (n = 734), Eastern (n = 634), Western (n = 818), Southern (n = 981), and North-Eastern (n = 117).

Mean domain scores varied across regions [Table 8; Figures 1 (a-c)]: knowledge ranged from 2.18 (North-Eastern) to 2.38 (Southern) out of 4, attitude from 0.73 (North-Eastern) to 1.00 (Western) out of 4, and practice from 43.7% (Northern) to 47.5% (Western). One-way ANOVA across the six regions indicated that differences in knowledge (F = 1.78, p = 0.114) and practice (F = 1.35, p = 0.241) did not reach statistical significance, but the difference in attitude was significant (F = 2.56, p = 0.026), driven by the North-Eastern region scoring consistently lower than the other five regions across nearly every individual attitude item. Effect sizes remained small for all three domains (η^2^ = 0.002-0.003), indicating that region explains only a modest share of variance even where the difference is statistically detectable.

**Table 8:**
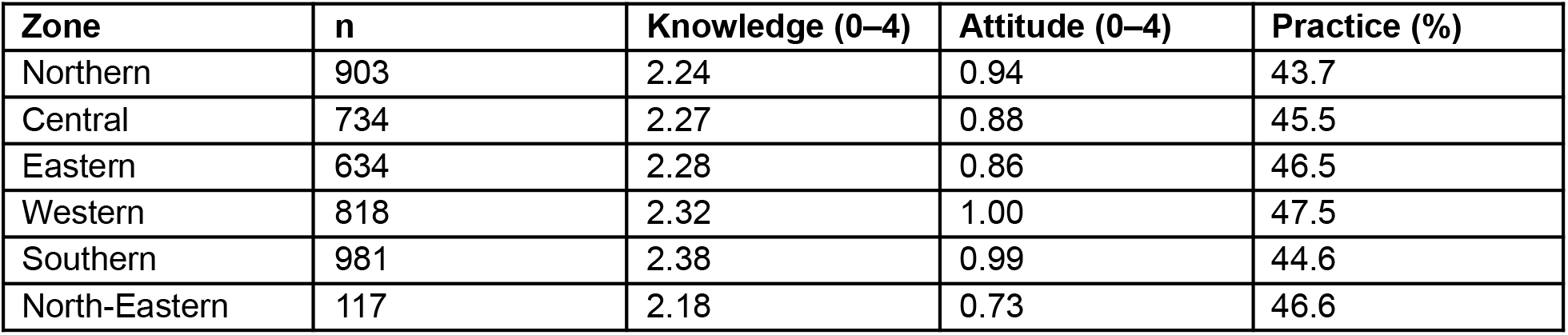
Mean knowledge, attitude and practice scores by region (n = 4,187)

| Zone | n | Knowledge (0–4) | Attitude (0–4) | Practice (%) |
| --- | --- | --- | --- | --- |
| Northern | 903 | 2.24 | 0.94 | 43.7 |
| Central | 734 | 2.27 | 0.88 | 45.5 |
| Eastern | 634 | 2.28 | 0.86 | 46.5 |
| Western | 818 | 2.32 | 1.00 | 47.5 |
| Southern | 981 | 2.38 | 0.99 | 44.6 |
| North-Eastern | 117 | 2.18 | 0.73 | 46.6 |

**Figure 1:**
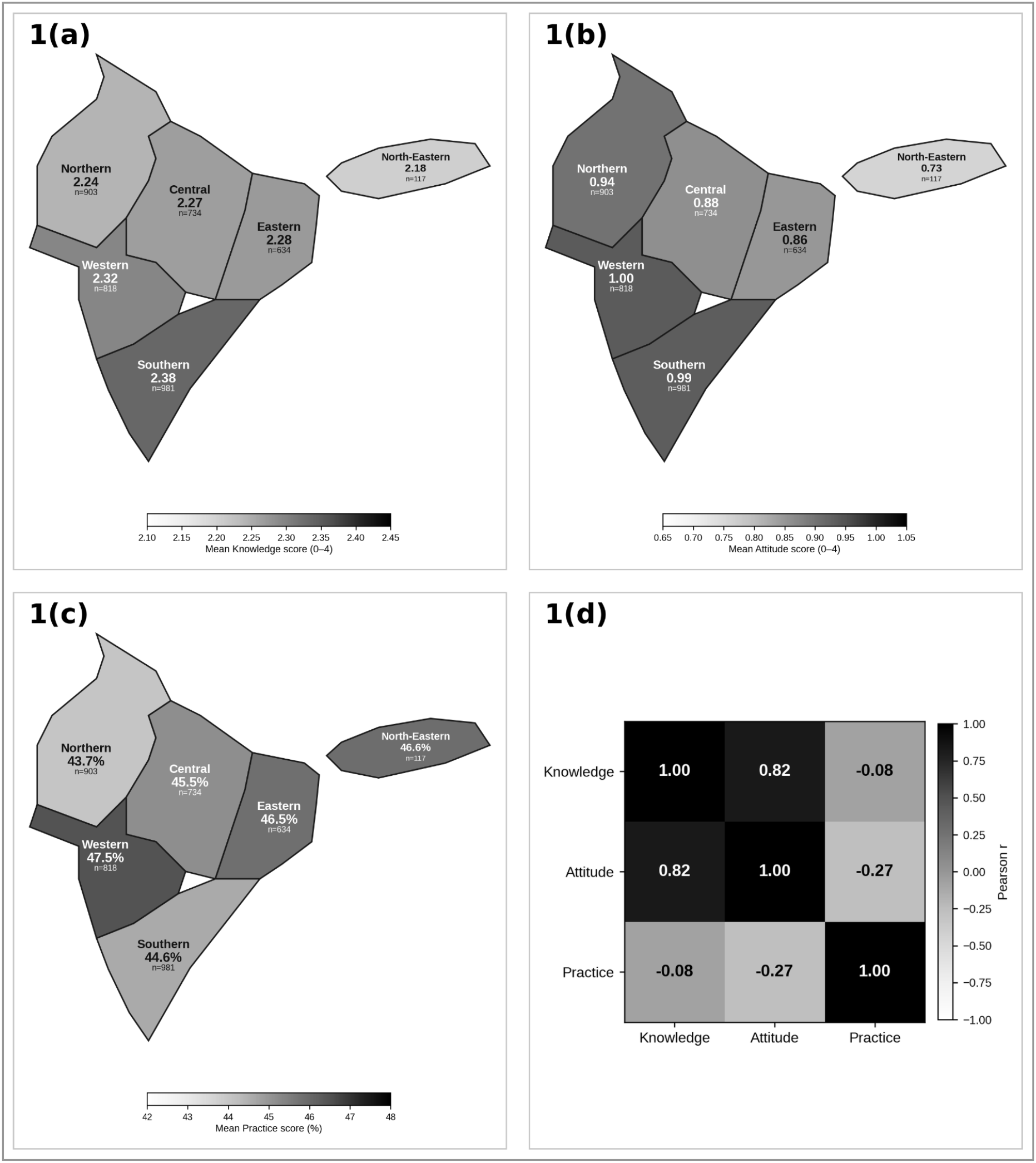
1(a) Mean knowledge score (0-4) by region. 1(b) Mean attitude score (0-4) by region. 1(c) Mean practice score (%) by region. 1(d) Region-level correlation between mean knowledge, attitude and practice (descriptive/exploratory; n = 6 regions, not respondent level).====Note: Regions follow the Ministry of Home Affairs Zonal Councils (States Reorganisation Act, 1956) for the five statutory zones, plus the North-Eastern region (North Eastern Council Act, 1971, including Sikkim), shown alongside rather than excluded. Zonal boundaries are schematic and not derived from surveyed cartographic data. Respondents per region: Northern n=903, Central n=734, Eastern n=634, Western n=818, Southern n=981, North-Eastern n=117

With regards to the regional distribution, regions with more accurate knowledge also tended to hold more favourable attitudes (Pearson r = 0.82 across the six regions). Practice scores were weakly and negatively correlated with both knowledge (r = −0.08) and attitude (r = −0.27) at this aggregate level (Figure 1d). These correlations are descriptive rather than inferential given the small number of units (n = 6). The pattern is addressed further in the Discussion.

Item-level analysis identified a broadly consistent pattern of best- and worst-understood concepts across the six regions (Figure 2). Recognition of common misuse behaviours (skipping doses, stopping early, self-prescribing, or reusing leftover antibiotics) was the best-performing item nationally (76-79% correct across regions), whereas awareness of having had AMR explained directly by a doctor was consistently the weakest knowledge item, ranging from 30% in the North-Eastern region to 49% in the Southern region. Among attitude items, the belief that antibiotics help resolve a cough or cold was the most resistant misconception in every region (12-16% correctly rejecting it). The North-Eastern region scored lowest of all six regions on three of the four attitude items, though not on A4 (reuse leftover antibiotics), where Central recorded the lowest score; North-Eastern also recorded the lowest score on the AMR-explained-by-doctor knowledge item.

**Figure 2:**
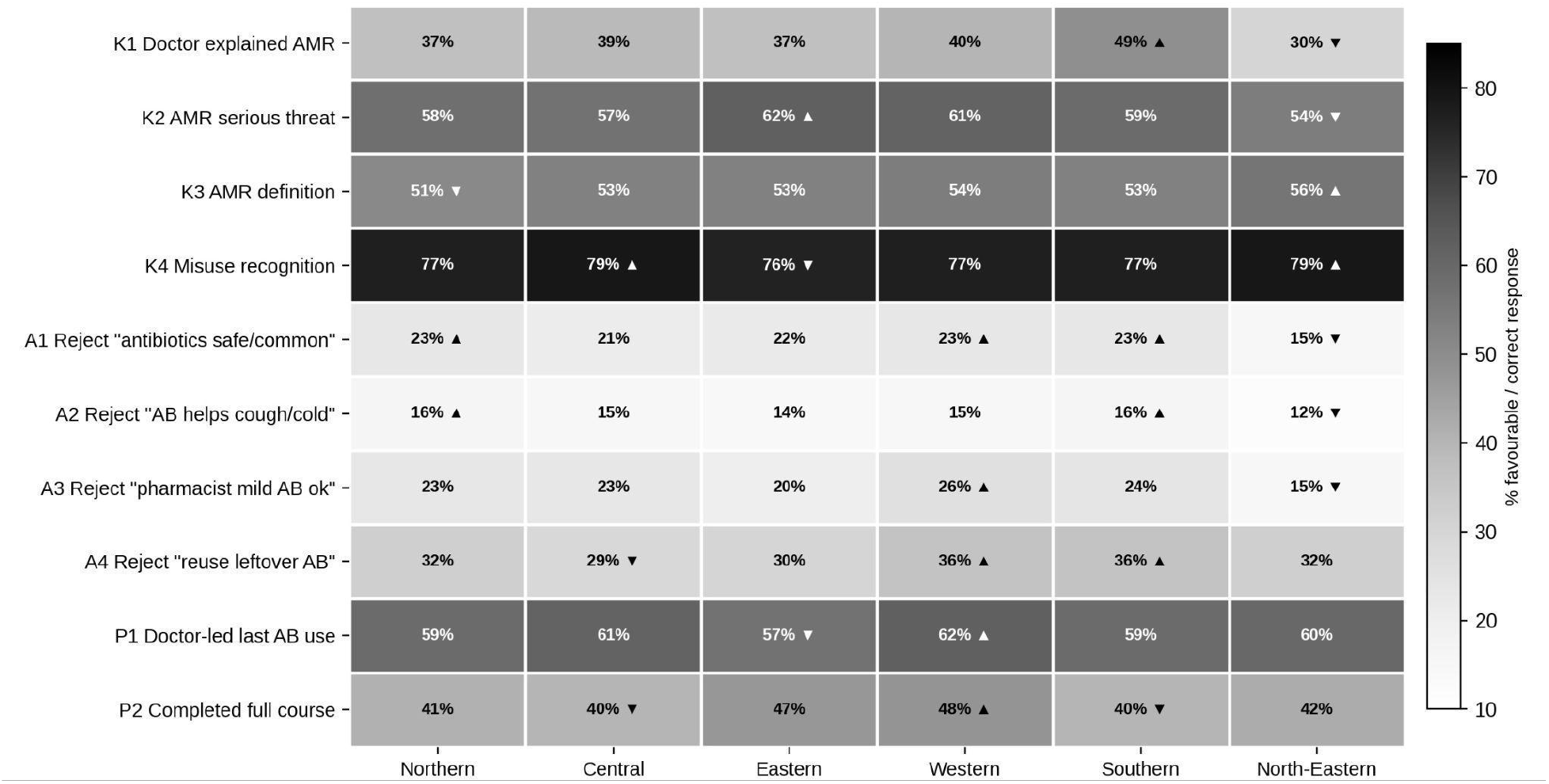
Best- and worst-answered knowledge, attitude and practice items, by region.====Note: Item wording abbreviated; n = 4,187. ▲ = region with high, ▼ = region with low % favourable response for that item.

### High-level Physician survey findings

Of 779 physician responses recorded, 256 were fully completed and formed the analytic sample. Respondents were predominantly general practitioners (n=127; 49.6%), followed by MD/MS/DNB specialists (n=78; 30.5%), super-specialists (n=22; 8.6%), dentists (n=9; 3.5%), and others (n=20; 7.8%). Practice settings were private hospital (n=106; 41.4%), private clinic (n=94; 36.7%), other (n=30; 11.7%), government (n=25; 9.8%), and academic (n=1; 0.4%). Years of clinical experience were evenly distributed across 1-5 years (n=86; 33.6%), 6-10 years (n=80; 31.3%), and 10+ years (n=90; 35.2%). Respondents practised in Tier 1 (n=110; 43.0%), Tier 2 (n=97; 37.9%), Tier 3 (n=33; 12.9%), and rural (n=16; 6.3%) settings.

### Comparative Highlights: Patients versus Physicians

We intentionally designed the two survey instruments in a way that several items in the two questionnaires assessed closely related or near-identical constructs from opposite ends of the same clinical encounter, to validate similar concepts across different perspectives. The comparisons are presented in Table 9. These are approached as an exploratory, hypothesis-generating triangulation across independently sampled respondent groups rather than a matched-pairs analysis, and are addressed further in the Discussion section.

**Table 9:**
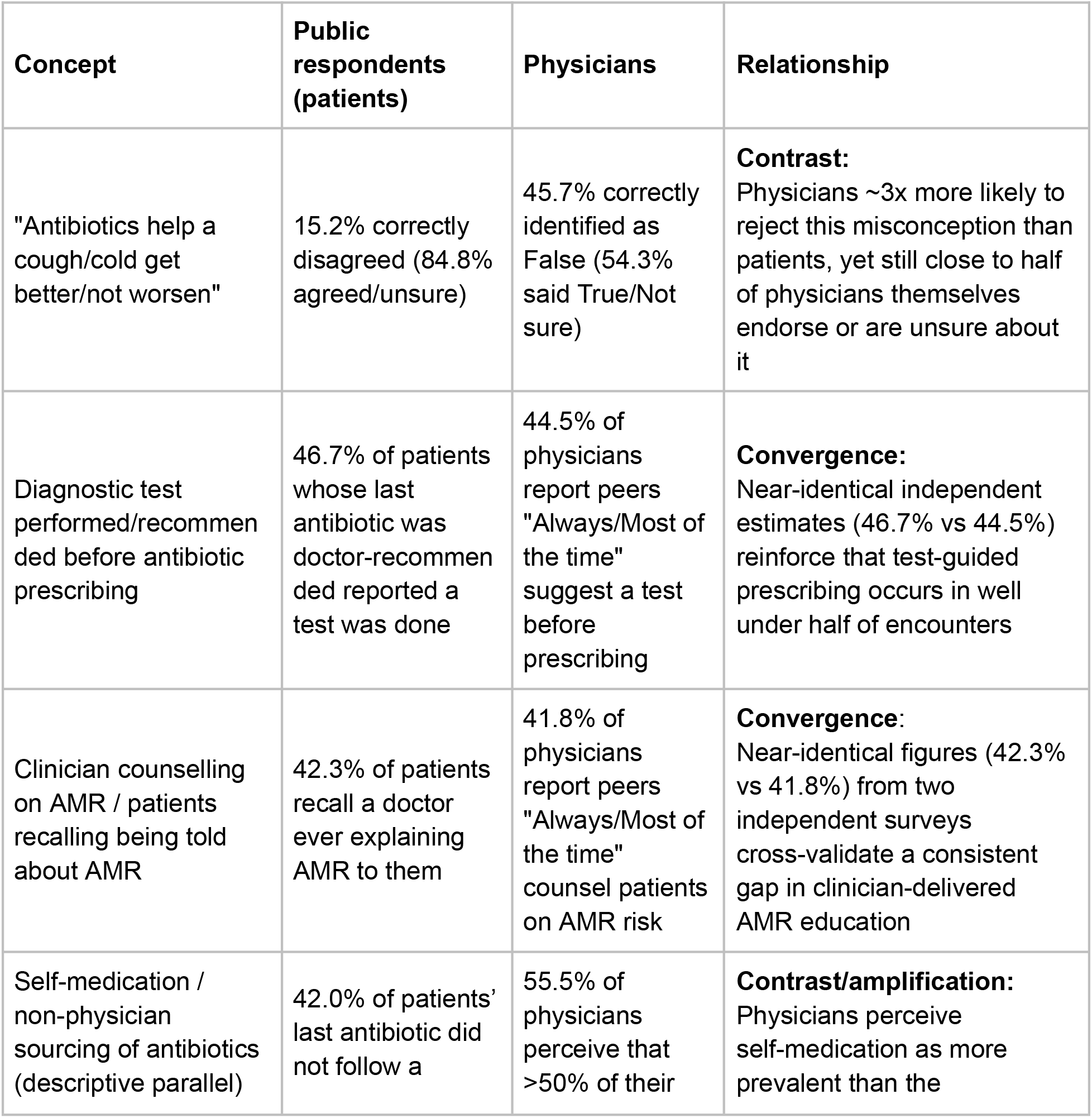
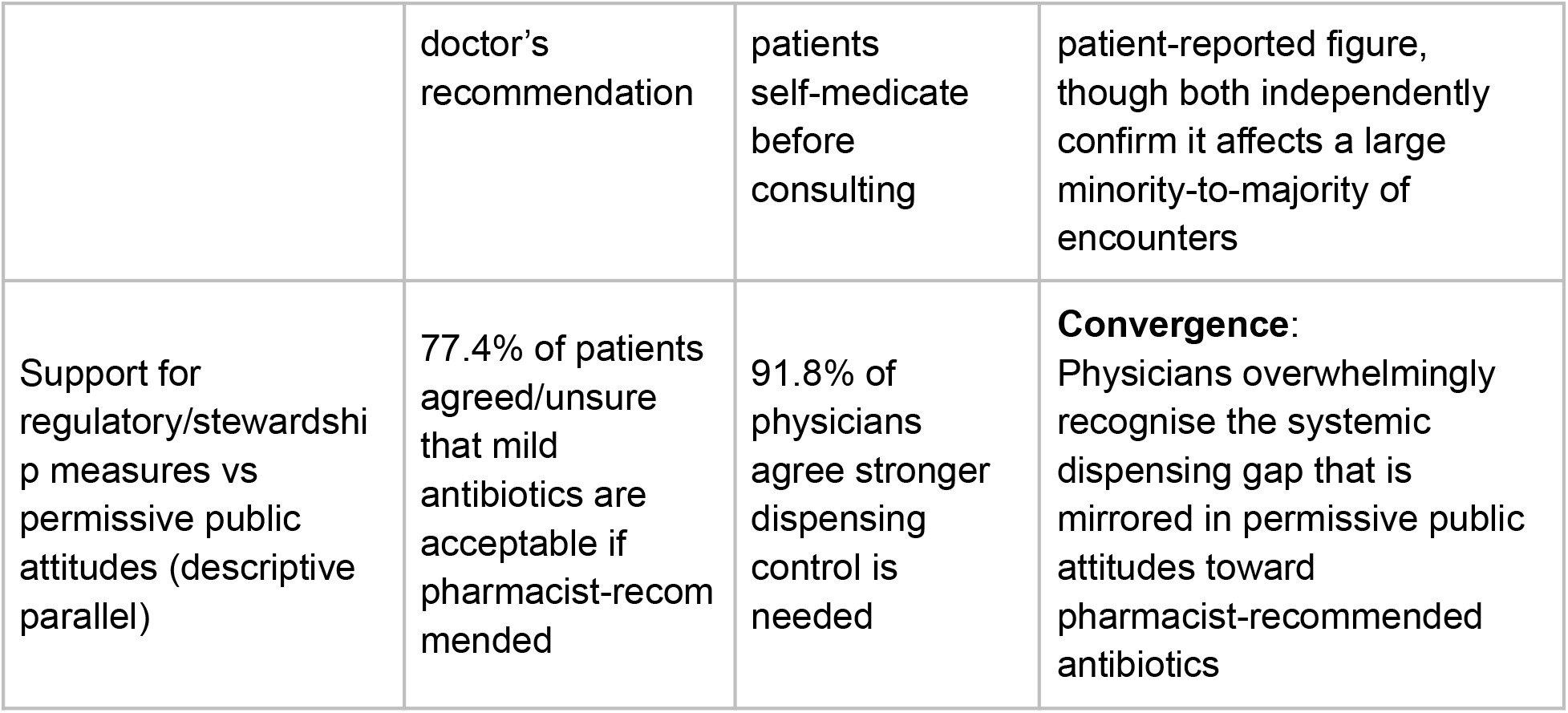
Comparative highlights across the parallel general public (patient) and physician surveys.

We had also asked the physicians to describe, in their own words, the main challenges they face with patients regarding antibiotic use (open-ended item; n = 129 responses), physicians most frequently cited concerns about resistance development in general (30%), followed by self-medication and easy over-the-counter access (12%), poor adherence to prescribed course or duration (12%), and reluctance or cost barriers to diagnostic testing before prescribing (9%) (Figure 3).

**Figure 3:**
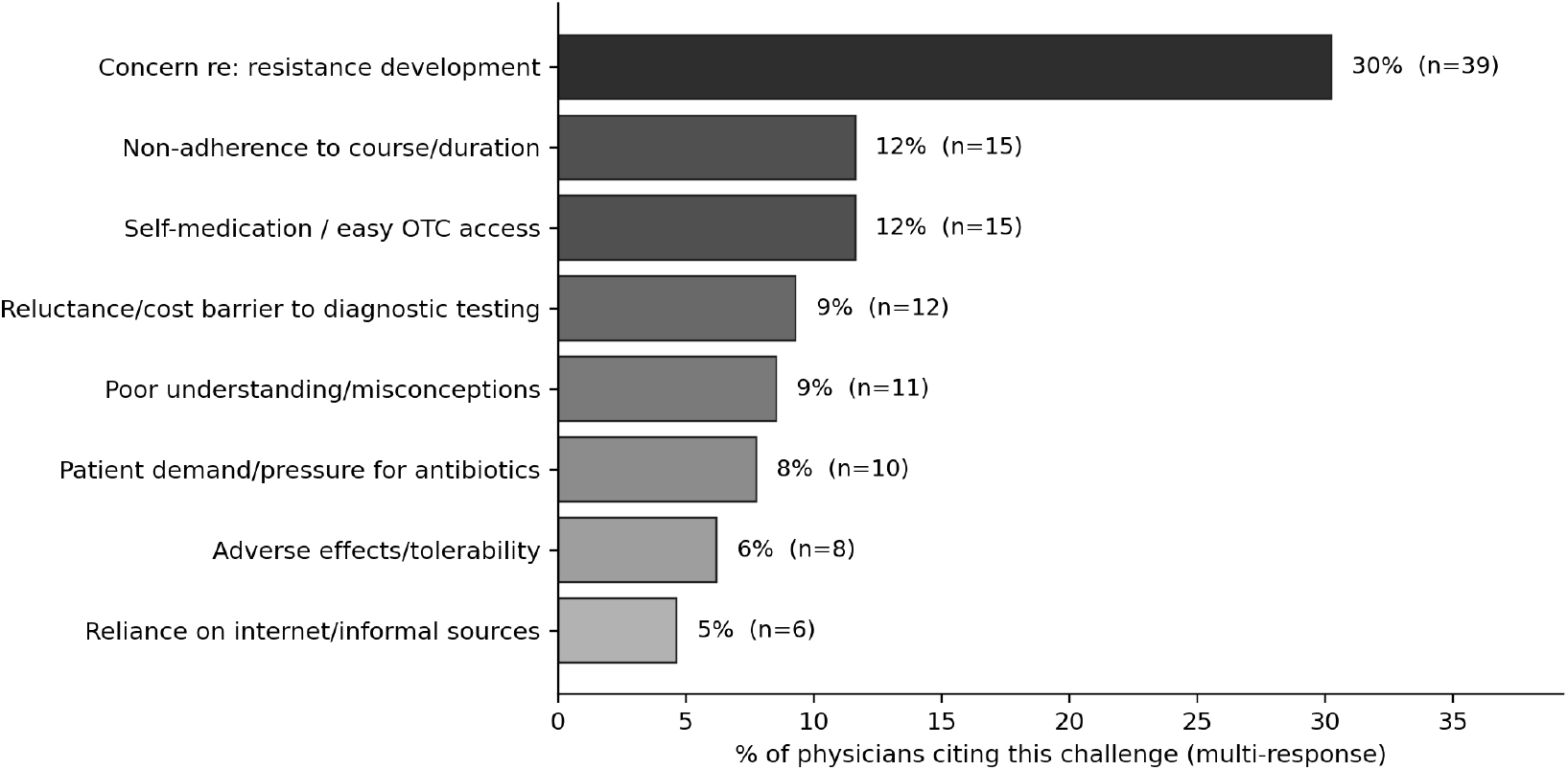
Physician-reported challenges with patients around antibiotic use, thematically coded from open-ended responses (n = 129)====Note: This is multi-label thematic coding, a response could be coded under more than one theme. For example, a single physician’s answer mentioning both cost barriers and self-medication gets counted in both bars. Hence, bar-level n is summing to less than 129, and percentages summing to less than 100%.

## Discussion

This study is among the first to describe knowledge, attitudes and practices around antibiotic use and resistance across India through a nationwide digital healthcare platform, and to examine how they vary by age, gender, city tier and region. Earlier Indian studies mostly described an awareness-practice gap, and our findings suggest a somewhat different picture. As measured here, attitudes were the weakest domain (23% favourable, against 57% for knowledge and 45% for practice), and knowledge and attitude scores moved together across the six regions (r = 0.82, descriptive). Across regions, practice was unrelated to knowledge (r = −0.08). The data therefore do not fit a simple account in which knowledge fails to translate into practice. They fit better with misconceptions that sit mostly in the attitude domain, while practice may depend on other things, possibly with access-related factors.

Item-level results sharpen this picture. Respondents were considerably better at recognising obvious misuse–self-medication, reusing leftover antibiotics–than at identifying why these behaviours are harmful, such as the distinction between bacterial and viral infection or the basic mechanism of resistance. This matches a pattern already visible in the Delhi[9], Karnataka[10], and central India[11] studies mentioned previously. The public seems to have absorbed the rules of appropriate antibiotic use without absorbing the reasoning behind them. What our data add is scale and precision, with a sample roughly three times larger than any single prior Indian KAP study, we can identify which specific concepts are weakest nationally, rather than inferring this from one city’s results.

The practice findings point toward the same structural explanation raised, but not tested, in earlier work. Close to half of respondents obtained their most recent antibiotic from a source other than a physician, consistent with the continued informal availability of antibiotics despite Schedule H1 restrictions. This was corroborated independently by our parallel physician survey. Over half of the surveyed physicians estimated that most patients self-medicate before a consultation. Furthermore, physicians rated their peers’ adherence to basic stewardship practices, such as documentation, pre-prescription testing, and resistance counselling, as low. Two independently sampled groups converging on the same picture makes it harder to dismiss this as self-report bias in either survey, and is consistent with inappropriate antibiotic use reflecting access pathways as well as knowledge gaps.

Another notable survey finding concerns what is possibly one of the most persistent misconceptions, that antibiotics treat colds and coughs. While physicians were three times as likely as patients to reject it (45.7% vs. 15.2%), nearly half of physicians still endorsed the myth or were unsure, showing this knowledge gap extends into medical practice and warrants targeted clinician education.

Demographic characteristics explained only a small share of the variation in KAP scores. Older respondents scored modestly higher across all three domains, which runs against the common assumption that younger, more digitally fluent users would show better platform-mediated health outcomes, plausibly because older adults have simply had more cumulative contact with physician-guided care. Females had a lower attitude score than males (p < 0.01) after adjusting for age and city tier. Knowledge score, too, was marginally lower (p < 0.1) and yet practice scores were not significantly different. Further research is needed to validate this relation and its associations. More notable than any demographic effect was that for a large share of respondents the last antibiotic was not physician-recommended, and that practice varied little with knowledge at the regional level. This points to access pathways as a plausible contributor to practice, a possibility these data cannot test, in line with calls to move from documenting what behaviours occur to explaining why they persist.

This pattern extended to how recently someone had used an antibiotic at all. Respondents reporting antibiotic use in the past 12 months scored significantly higher across all three domains than those reporting no recent use or uncertainty (Knowledge 2.54 vs. 2.15 vs. 1.90; Attitude 1.06 vs. 0.82 vs. 0.71; Practice 50.7% vs. 41.0% vs. 38.3%; all p < 0.001). This held most strongly for practice, consistent with the recall-bias rationale for the 12-month criterion, which applies most directly to items asking about a specific recent event and less to knowledge and attitude items concerning general beliefs. Recent personal experience with the care pathway, in other words, tracks stronger outcomes independently of demographics, reinforcing the same point above: what someone has actually done recently predicts more than what they officially know.

Geography adds a different angle. Prior work in this space has relied almost entirely on single-city or single-event cohorts.[4-6] Mapping our sample across six regions of India lets us check whether that picture holds nationally, and largely, it does: knowledge and practice varied only marginally by region, with no significant differences. Attitude was the exception, significantly lower in the North-Eastern region (F = 2.56, p = 0.026), consistent across nearly every attitude item, though the effect size was small (η^2^ = 0.003) and shouldn’t be read as a large disparity. Set against the Delhi and Karnataka findings, this suggests the awareness-practice paradox seen in individual cities isn’t local to those settings but holds nationally, at least among digitally engaged users. That argues for one national stewardship message over a portfolio of regional campaigns, with North-Eastern as the one targeted exception.

This study’s principal strengths lie in its scale and setting. At n = 4,883, it is among the largest single-platform KAP surveys of the Indian general public conducted to date, exceeding prior high-reach cohorts such as the 2022 central India mass-gathering study (n = 1,915) and the 2025 urban Delhi survey (n = 1,601), with respondents drawn from Tier 1, 2, and 3 cities rather than a single locality. By design, conditional branching within the practice domain ensured respondents were only asked questions relevant to their own most recent antibiotic-use experience, improving contextual validity. Integration within a platform respondents were already using for healthcare, rather than a dedicated field survey, further allowed behavioural data to be captured at scale from an actively engaged population.

These strengths come with corresponding limitations. The cross-sectional design cannot establish the direction of the associations observed between participant characteristics and KAP outcomes. Invitation through a digital, English-language platform likely selected for users who are more digitally literate, more comfortable in English, and plausibly more health-literate than the general population; consequently, the KAP levels reported here may represent an upper bound rather than a nationally representative sample, and off-platform or less digitally literate groups could plausibly show lower awareness and less favourable practice than observed in this sample. All outcome measures relied on self-report and are subject to recall and social-desirability bias. The questionnaire was assessed by face validation only; future versions should be validated through a Delphi process with subject experts. Also, a web-based convenience sample is likely to introduce selection bias toward more active platform users, who may exhibit different behaviors and attitudes than passive users. The use of convenience sampling through a web-based channel is also likely to have introduced selection bias. The practice composite itself, built from a branching item set, trades item-level comparability across respondents for clinical relevance, and assumes equal weighting across its constituent items, an assumption not independently verified. Finally, unmeasured variables including educational attainment, income, health literacy, prior healthcare experience, were not collected, and likely account for a meaningful share of the variance our regression models left unexplained.

Taken together, these findings argue for a specific and structural role for digital health platforms, not simply more awareness. Establishing a pan-India baseline of this kind is a prerequisite for transforming digital pharmacy ecosystems from passive supply chains into targeted intervention points under NAP-AMR 2.0, and three interventions follow directly from what this study identified as weakest. First, intelligent nudging: brief, interactive competency checks at the point of purchase, targeted at the concepts our data show are worst understood, the bacterial-versus-viral distinction chief among them, rather than generic AMR messaging. Second, a regional hotspot approach: our data provide a concrete first candidate for this, with the North-Eastern region showing significantly less favourable attitudes than the rest of the country, particularly around common antibiotic misconceptions, warranting priority stewardship content in that region specifically rather than a uniform national rollout. Third, repositioning the platform itself as a validation layer rather than a passive dispensing channel, a “digital stewardship protocol” in which prescription verification becomes a condition of fulfilment, shifting such platforms from just ePharmacies to active gatekeepers.

## Conclusion

This survey of 4,883 users of a national digital healthcare platform provides contemporary evidence on antibiotic knowledge, attitudes and practices in India. Attitudes, not knowledge, were the weakest domain, and for 42.0% of respondents the last antibiotic was not physician-recommended, a pattern the parallel physician survey also described. Knowledge and practice were broadly similar across regions, and demographic characteristics explained only a small share of the variation. These findings suggest that digital healthcare platforms could serve as intervention points for antimicrobial stewardship under NAP-AMR 2.0, although the cross-sectional design and the platform-user sample limit how far they generalise. Future research should examine how behavioural, socioeconomic and health-system factors interact to shape antibiotic use, and evaluate whether digitally delivered interventions improve stewardship outcomes.

## Data Availability

All data produced in the present study are available upon reasonable request to the authors

## Declarations

### Ethics approval and consent to participate

Ethics approval for secondary analysis of the survey was obtained from Genebandhu Independent Ethics Committee (reference number ECG013/2026). Under the platform’s Terms and Conditions, users consent to receive health-related communications and participate in platform-based surveys; completion of the questionnaire was therefore considered to imply informed consent. Participation was entirely voluntary. The questionnaire collected no names or contact details, and results are reported in aggregate only.

### Consent to publication

Not applicable

### Availability of Data

The datasets generated and analysed during this study are available from the corresponding author on reasonable request. The questionnaire and scoring scheme are provided as Supplementary Tables 2 and 3.

### Competing Interests

The study was conducted among users of the Tata 1mg platform, which served as the channel for inviting participants and the source of survey responses. ZA is an intern at Tata 1mg, and SM and RS are employed by Tata 1mg; JK and TS are independent contributors. No author received compensation linked to the study outcome. The study was designed, conducted and interpreted by the Medical Affairs research team and independent contributors; business and commercial teams had no role in study design, data collection, analysis, interpretation, or the decision to submit the manuscript for publication.

### Funding

The authors received no specific funding or external financial sponsorship for this work. The survey platform, infrastructure and staff time of ZA, SM and RS were provided by Tata 1mg; no other payment or services were received from a third party.

### Authors’ contributions

ZA (Zoya Agarwal), SM (Swati Mishra), RS (Rajeev Sharma), JK (Jasmine Kaur), and TS (Tavpritesh Sethi) compiled the study architecture. ZA, SM, and RS conceived and designed the study. SM was responsible for platform-related implementation and data acquisition. ZA and SM performed the statistical analysis, interpreted the data, and drafted the manuscript. JK provided additional inputs on interpretation of findings. RS, JK, and TS critically revised the manuscript for important intellectual content. All authors read and approved the final manuscript.

## Acknowledgments

The authors would like to sincerely thank Monalisa Deka and Nitin Negi for helping coordinate the initial stages of the survey.

## Notes

### Author Declarations

Genebandhu Independent Ethics Committee (reference number ECG013/2026) gave ethical approval for this work

