## Supplementary figures and tables for "Knowledge, Attitudes, and Practices Regarding Antimicrobial Resistance Among Users of an Indian Digital Healthcare Platform: A Cross-sectional Online Survey": Preprint AMR KAP in India_Supplementary Figures & tables.docx

**Supplementary Table 1: Comparison of study questionnaire items with the WHO multi-country public awareness survey**

| **Study item** | **Domain** | **Study questionnaire item** | **Corresponding WHO item** | **Status** | **Reason for change** |
| --- | --- | --- | --- | --- | --- |
| Q1 | Demographics | Which age bracket do you fall into? (18-34 years; 35-64 years; ≥65 years) | S2: And what age are you now? (16-18; 19-24; 25-34; 35-44; 45-54; 55-64; 65+) | Modified | Age bands widened and reduced from seven to three categories |
| Q2 | Demographics | Please select your gender. (Male; Female; Prefer not to mention) | S1: To begin, please record gender. (Male; Female) | Modified | "Prefer not to mention" option added |
| Q3 | Demographics | Which city or town do you currently live in? (Free text) | S3: In which of these regions do you live? (Single code, adapted per country) | Modified | Changed from closed-code region selection to free-text city entry, for finer-grained geographic mapping |
| Q4 | Practice | Have you taken any antibiotics in the last 12 months? (Yes; No; Don't remember) | Q1: When did you last take antibiotics? (In the last month; In the last 6 months; In the last year; More than a year ago; Never; Can't remember) | Modified | Recency bands collapsed into a single 12-month exposure window; to support recall accuracy for practice-domain items and capture respondents' most recent antibiotic-use experience |
| Q5 | Practice | Who recommended the last antibiotic you used? (Doctor; Pharmacist; Self; Family/Friends; Previous prescription) | Q2: On that occasion, did you get the antibiotics (or a prescription for them) from a doctor or nurse? (Yes; No; Can't remember) / Q4: On that occasion, where did you get the antibiotics? (Medical store or pharmacy; Stall or hawker; The internet; Friend or family member; I had them saved up from a previous time; Somewhere/someone else; Can't remember) | Modified | Two WHO items (source confirmation and place of acquisition) combined into one recommendation-source item; also used as conditional branching |
| Q6 | Practice | Why did you choose not to consult a doctor? (Mild symptoms; Doctor consultation is costly; Lack of time; Trust my pharmacist; Others) | Not in WHO questionnaire | Newly added | Added to capture reasons for non-doctor pathways, relevant to the Indian retail pharmacy and self-medication context |
| Q7 | Practice | Did you finish the entire antibiotic course? (Always; Most of the times; Sometimes; Never) | Q5: When do you think you should you stop taking antibiotics once you've begun treatment? (When you feel better; When you've taken all of the antibiotics as directed; Don't know) | Modified | Converted from a belief/knowledge item (what one should do) to a self-reported practice item (what one actually did) |
| Q8 | Practice | Did your doctor recommend any diagnostic tests before prescribing the antibiotic? (Yes; No; Not sure) | Not in WHO questionnaire | Newly added | Added to capture diagnostic-testing practice, not covered in the WHO instrument |
| Q9 | Attitude | Antibiotics are safe and effective, so they can be used commonly. (Agree; Disagree; Not sure) | Not in WHO questionnaire | Newly added | Added to capture a common logical slippage: people often correctly believe antibiotics are safe and effective when properly indicated, but extend that correct belief into an incorrect inference that frequent or casual use is therefore acceptable. |
| Q10 | Attitude | Taking antibiotics can stop a cough or cold from getting worse. (Agree; Disagree; Not sure) | Q8: Do you think these conditions can be treated with antibiotics? - item: Cold and flu | Modified | One condition drawn out of WHO's 12-condition checklist and reworded as a standalone attitude statement; remaining 11 conditions dropped |
| Q11 | Attitude | Some mild antibiotics are okay to be taken if a pharmacist recommends them. (Agree; Disagree; Not sure) | Not in WHO questionnaire | Newly added | New item reflecting pharmacist-led dispensing, common in the Indian retail pharmacy setting |
| Q12 | Attitude | Leftover or unused antibiotics can be taken for similar illness in future. (Agree; Disagree; Not sure) | Q12, statement 3: People should not keep antibiotics and use them later for other illnesses (5-point agree/disagree scale) | Modified | Reworded from a prescriptive action statement to a personal-belief statement; scale collapsed from 5-point to agree/disagree/not sure |
| Q13 | Knowledge | Has your doctor ever explained antibiotic resistance (AMR) to you? (Yes; No; Don't remember) | Not in WHO questionnaire | Newly added | Added to capture source of knowledge on AMR specifically; loosely related to WHO's source-of-awareness question (Q10 series) but structurally different |
| Q14 | Knowledge | Antibiotic resistance is a growing serious threat for all of us. (True; False; Not sure) | Q13, statement 1: Antibiotic resistance is one of the biggest problems the world faces (5-point agree/disagree scale) | Modified | Reworded as a true/false knowledge-style item; scale collapsed from 5-point to true/false/not sure |
| Q15 | Knowledge | Antibiotic resistance occurs when antibiotics fail to treat infections. (True; False; Not sure) | Q11, statement 1: Antibiotic resistance occurs when your body becomes resistant to antibiotics and they no longer work as well (True; False; Don't know) | Modified | WHO's statement tests the common misconception (body becomes resistant); reworded here to state the correct definition directly |
| Q16 | Knowledge | Which of these counts as misuse of antibiotics? Multiple response (Skipping doses; Stopping the course early; Taking antibiotics without a doctor's prescription; Using leftover antibiotics; None of the above) | Not in WHO questionnaire as a single item - draws on Q5 (stopping early) and Q6 (leftover antibiotics) | Modified | Several separate WHO concepts (course completion, self-prescription, leftover use) consolidated into one multi-response misuse-recognition item, for brevity |

**Supplementary Table 2: Survey questionnaire and scoring scheme for the general public**

| Section | Q. No. | Survey Question | Response Options | Scoring |
| --- | --- | --- | --- | --- |
| Demographics* | Q1 | Which age bracket do you fall into? | 18–34 years; 35–64 years; ≥65 years | Not scored |
|  | Q2 | Please select your gender. | Male; Female; Prefer not to mention | Not scored |
|  | Q3 | Which city or town do you currently live in? | Free text | Not scored |
| Practice | Q4 | Have you taken any antibiotics in the last 12 months? | Yes; No; Don't remember | Not scored (Exposure variable) |
|  | Q5** | Who recommended the last antibiotic you used? | Doctor; Pharmacist; Self; Family/Friends; Previous prescription | Doctor = 1, all other responses = 0. (Also acts as skip logic question) |
|  | Q6 | Why did you choose not to consult a doctor? | Mild symptoms; Doctor consultation is costly; Lack of time; Trust my pharmacist; Others | Not scored |
|  | Q7 | Did you finish the entire antibiotic course? | Always; Most of the times; Sometimes; Never | Always = 1; all other responses = 0 |
|  | Q8 | Did your doctor recommend any diagnostic tests before prescribing the antibiotic? | Yes; No; Not sure | Yes = 1; No/Not sure = 0 |
| Attitude | Q9 | Antibiotics are safe and effective, so they can be used commonly. | Agree; Disagree; Not sure | Disagree = 1; Agree/Not sure = 0 |
|  | Q10 | Taking antibiotics can stop a cough or cold from getting worse. | Agree; Disagree; Not sure | Disagree = 1; Agree/Not sure = 0 |
|  | Q11 | Some mild antibiotics are okay to be taken if a pharmacist recommends them. | Agree; Disagree; Not sure | Disagree = 1; Agree/Not sure = 0 |
|  | Q12 | Leftover or unused antibiotics can be taken for similar illness in future. | Agree; Disagree; Not sure | Disagree = 1; Agree/Not sure = 0 |
| Knowledge | Q13 | Has your doctor ever explained antibiotic resistance (AMR) to you? | Yes; No; Don't remember | Yes = 1; No/Don't remember = 0 |
|  | Q14 | Antibiotic resistance is a growing serious threat for all of us. | True; False; Not sure | True = 1; False/Not sure = 0 |
|  | Q15 | Antibiotic resistance occurs when antibiotics fail to treat infections. | True; False; Not sure | True = 1; False/Not sure = 0 |
|  | Q16 | Which of these counts as misuse of antibiotics? (Multiple response) | Skipping doses; Stopping the course early; Taking antibiotics without a doctor's prescription; Using leftover antibiotics; None of the above | Selection of at least one correct option (a–d) = 1; None of the above or no correct option = 0 |

*Questions Q1-Q3 collected demographic characteristics and were not included in the scoring, and were not mandatory.
**If the respondent selects "Doctor", the survey skips directly to Questions 7 and 8. For all other responses, Questions 6 and 7 are displayed.

**Supplementary Table 3: Scoring summary for KAP survey questionnaire used for general public**

| **Domain** | **Items** | **Score Range** |
| --- | --- | --- |
| Practice | 3 | 0–3 |
| Attitude | 4 | 0–4 |
| Knowledge | 4 | 0–4 |

**Figure 1: Participation funnel for the AMR KAP survey in general public**
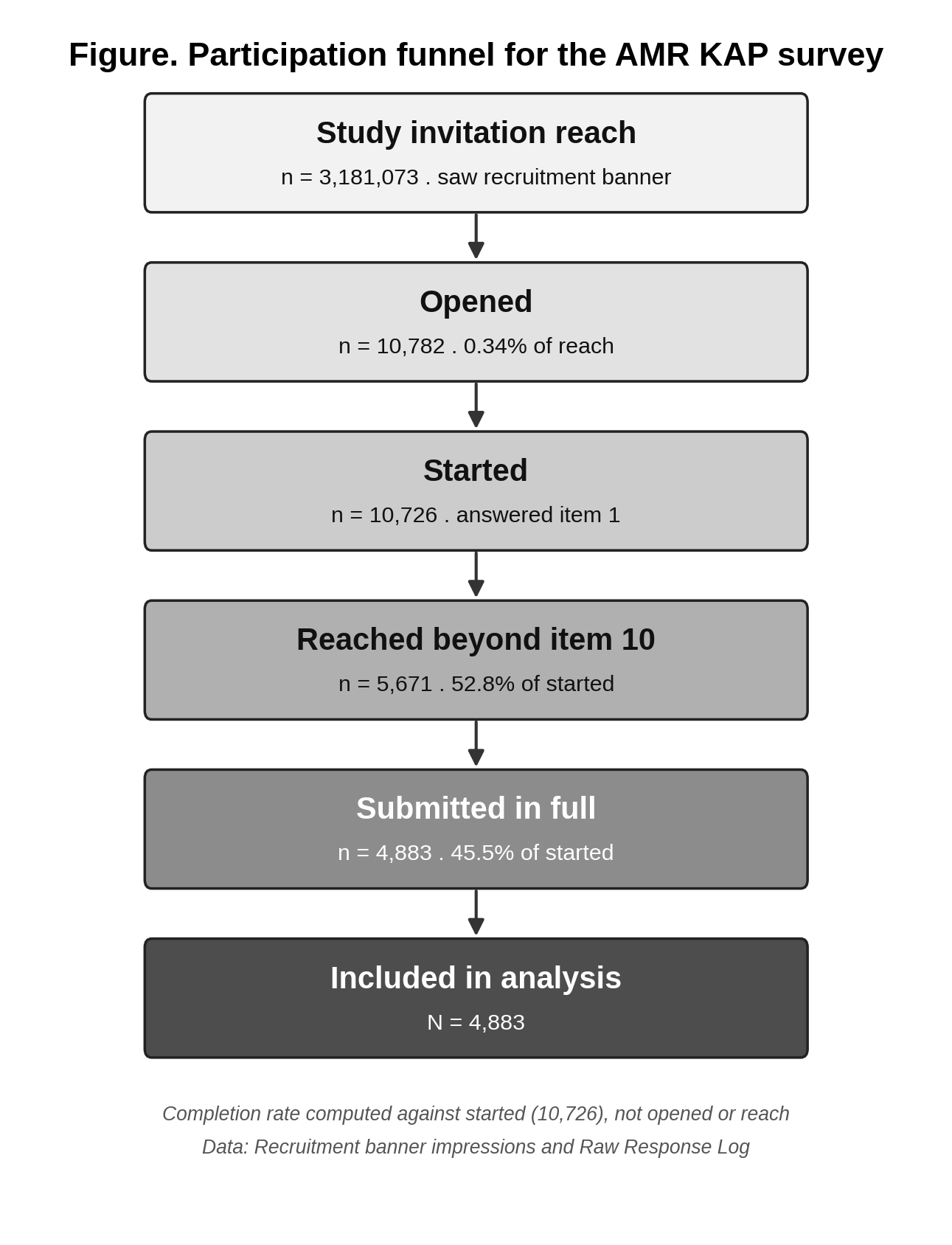

*Note: Completion rate has been computed against survey started (10,726).*

**Supplementary Table 4: Survey questionnaire and scoring scheme for the physicians**

| Section 1: Background |  |  |  |
| --- | --- | --- | --- |
| **Question** | **Response** | **Score** |  |
| 1. Please indicate your medical qualification or specialty | General Practitioner (MBBS / GP) | Not scored |  |
|  | Dentist (BDS/ MDS) | Not scored |  |
|  | MD/ MS/ DNB | Not scored |  |
|  | Super specialization (DM / MCh / Fellowship) | Not scored |  |
|  | Others | Not scored |  |
| 2. What is your practice setting? | Government | Not scored |  |
|  | Private hospital | Not scored |  |
|  | Private clinic | Not scored |  |
|  | Academics | Not scored |  |
|  | Others | Not scored |  |
| 3. How many years of clinical experience do you have? | 1-5 years | Not scored |  |
|  | 6-10 years | Not scored |  |
|  | 10+ years | Not scored |  |
| 4. What is your current place of practice? | Tier 1 | Not scored |  |
|  | Tier 2 | Not scored |  |
|  | Tier 3 | Not scored |  |
|  | Rural | Not scored |  |
| Section 2: Knowledge |  |  |  |
| **Question** | **Response** | **Score** | **Rationale** |
| 5. Stopping antibiotics before completing the prescribed course can lead to antibiotic resistance | TRUE | 1 | Correct understanding of antibiotic resistance development |
|  | False / Not sure | 0 | Incorrect or uncertain knowledge |
| 6. AMR can develop even when antibiotics are used appropriately | TRUE | 1 | Recognizes that resistance can emerge despite appropriate use |
|  | False / Not sure | 0 | Incorrect or uncertain knowledge |
| 7. Antibiotic resistance can spread from one patient to another in healthcare settings | TRUE | 1 | Correct understanding of transmission of resistant organisms |
|  | False / Not sure | 0 | Incorrect or uncertain knowledge |
| 8. Antibiotics can stop a cough or cold from getting worse | FALSE | 1 | Correctly identifies that antibiotics are generally ineffective for viral illnesses |
|  | True / Not sure | 0 | Incorrect or uncertain knowledge |
| 9. Antibiotics help patients recover faster from a fever | FALSE | 1 | Correctly recognizes that fever alone does not indicate a need for antibiotics |
|  | True / Not sure | 0 | Incorrect or uncertain knowledge |
| 10. Unlike systemic use, topical antibiotics are not known to be a key driver of AMR | False* | 1 | Correctly recognizes that topical antibiotic use may contribute to AMR |
|  | True / Not sure | 0 | Incorrect or uncertain knowledge |
| 11. Clinical studies in India have identified bacteria harboring the mcr-1 and blaNDM-1 resistance genes | TRUE | 1 | Demonstrates awareness of important resistance mechanisms reported in India |
|  | False / Not sure | 0 | Incorrect or uncertain knowledge |
| **Question** | **Response** | **Score** | **Rationale** |
| 12. Patients expect antibiotics even when they are not needed | Agree | 1 | Recognizes a known driver of inappropriate antibiotic use |
|  | Disagree / Not sure | 0 | Does not recognize or is uncertain about this driver |
| 13. Uncertainty in diagnosis often causes doctors to prescribe antibiotics unnecessarily | Agree | 1 | Acknowledges a documented contributor to antibiotic overprescribing |
|  | Disagree / Not sure | 0 | Does not recognize or is uncertain about this contributor |
| 14. Stronger control over antibiotic dispensing is needed to reduce resistance | Agree | 1 | Supports antimicrobial stewardship and regulatory interventions |
|  | Disagree / Not sure | 0 | Does not support or is uncertain about stewardship measures |
| 15. Doctors in India receive sufficient training on antibiotic stewardship to prescribe responsibly and prevent AMR | Not scored* | – | Perception-based item with no objectively correct response |
| **Question** | **Response** | **Score** | **Rationale** |
| 16. In your experience, what proportion of your patients self-medicate with antibiotics before consulting you? | Not scored* | – | Reflects patient behaviour rather than physician practice |
| 17. Among your peers, how commonly are the indication, dose, and duration recorded for antibiotic prescriptions? | Always / Most of the times | 1 | Reflects stewardship-concordant prescribing and documentation practices |
|  | Sometimes / Never | 0 | Reflects suboptimal documentation practices |
| 18. From what you observe in your practice settings, how often do clinicians suggest diagnostic tests before prescribing antibiotics? | Always / Most of the times | 1 | Reflects diagnostic stewardship and evidence-based prescribing |
|  | Sometimes / Never | 0 | Reflects limited use of diagnostics prior to antibiotic prescribing |
| 19. Within your clinical community, how often do clinicians counsel their patients about the risks of antibiotic resistance? | Always / Most of the times | 1 | Reflects stewardship-supportive patient education practices |
|  | Sometimes / Never | 0 | Reflects inadequate AMR counselling practices |
| 20. What are the main challenges you face with patients related to antibiotic use? | Free text; Not scored* |  | Experience-based item with no objectively correct response |
| *Excluded from composite score calculations and analysed descriptively because the item assesses perceptions or contextual observations rather than stewardship-concordant knowledge, attitudes, or practices. |  |  |  |

- Inclusion criteria
  Registered Tata 1mg users aged 18 years or older
  residing in India.
- Exclusion criteria were
  Duplicate submissions, incomplete questionnaires (defined as responses that did not reach the final item), any malformed responses (for example, only a single logged answer with no other data, consistent with a data-logging artefact rather than genuine survey engagement).

**Supplementary Table 5: Summary of geographical mapping of cities**

| **Metric** | **Count** | **% (of 4,883)** |
| --- | --- | --- |
| Total completed respondents (N) | 4883 | 100 |
| Recognized to a valid place (final) | 4195 | 85.9 |
| Still unclassified / Invalid response / international | 688 | 14.1 |

**Supplementary Table 6: State-wise breakdown of the number of respondents by region**

| **State** | **Count** |
| --- | --- |
| ***Northern (n = 903)*** |  |
| Delhi | 327 |
| Haryana | 225 |
| Rajasthan | 156 |
| Punjab | 114 |
| Jammu and Kashmir | 41 |
| Himachal Pradesh | 26 |
| Chandigarh | 14 |
| ***Central (n = 734)*** |  |
| Uttar Pradesh | 510 |
| Madhya Pradesh | 123 |
| Uttarakhand | 55 |
| Chhattisgarh | 46 |
| ***Eastern (n = 634)*** |  |
| West Bengal | 247 |
| Bihar | 198 |
| Odisha | 107 |
| Jharkhand | 82 |
| ***Western (n = 818)*** |  |
| Maharashtra | 664 |
| Gujarat | 131 |
| Goa | 18 |
| Dadra and Nagar Haveli and Daman and Diu | 5 |
| ***Southern (n = 981)*** |  |
| Karnataka | 350 |
| Tamil Nadu | 232 |
| Telangana | 208 |
| Andhra Pradesh | 107 |
| Kerala | 77 |
| Puducherry | 7 |
| ***North-Eastern (n = 117)*** |  |
| Assam | 66 |
| Meghalaya | 20 |
| Manipur | 7 |
| Sikkim | 7 |
| Arunachal Pradesh | 6 |
| Mizoram | 5 |
| Tripura | 4 |
| Nagaland | 2 |
| ***Unclassified (UT)*** |  |
| Andaman and Nicobar Islands | 8 |
